# Timing of Initiation and Efficacy of Dual Antiplatelet Therapy in Minor Stroke or High-Risk TIA

**DOI:** 10.1101/2025.08.11.25333465

**Authors:** Jaemin Shin, Keon-Joo Lee, Chi Kyung Kim, Kyumgmi Oh, Do Yeon Kim, Beom Joon Kim, Moon-Ku Han, Hyunsoo Kim, Joon-Tae Kim, Kang-Ho Choi, Dong-Ick Shin, Kyu Sun Yum, Jae-Kwan Cha, Dae-Hyun Kim, Dong-Eog Kim, Dong-Seok Gwak, Jong-Moo Park, Dongwhane Lee, Kyusik Kang, Soo Joo Lee, Jae Guk Kim, Kyung-Ho Yu, Mi-Sun Oh, Minwoo Lee, Keun-Sik Hong, Yong-Jin Cho, Hong-Kyun Park, Jay Chol Choi, Joong-Goo Kim, Tai Hwan Park, Sang-Soon Park, Jee-Hyun Kwon, Wook-Joo Kim, Jun Lee, Doo Hyuk Kwon, Sung-Il Sohn, Jeong-Ho Hong, Hyungjong Park, Kyungbok Lee, Jeong-Yoon Lee, Kwang-Yeol Park, Hae-Bong Jeong, Chulho Kim, Sang-Hwa Lee, Sung Hyuk Heo, Ho Geol Woo, Ji Sung Lee, Juneyoung Lee, Hee-Joon Bae, the CRCS-K-NIH investigators

## Abstract

**Background:** Dual antiplatelet therapy (DAPT) with aspirin and clopidogrel is recommended within 24 hours for patients with minor ischemic stroke or high-risk transient ischemic attack (TIA). However, the optimal timing for initiating DAPT in real-world practice remains unclear.

**Methods:** We conducted a secondary analysis of the Clinical Research Center for Stroke–Korea National Institute of Health (CRCS-K-NIH) registry, a prospective multicenter cohort involving 20 stroke centers across South Korea between January 2011 and April 2023. A total of 41,530 patients with minor non-cardioembolic ischemic stroke (NIHSS ≤5) or high-risk TIA who presented within 7 days of symptom onset were included. We evaluated outcomes based on in-hospital initiation of DAPT versus monotherapy (aspirin or clopidogrel alone). The primary outcome was a composite of recurrent stroke, myocardial infarction, and death within 90 days. Time-to-treatment effects were analyzed using Cox proportional hazards models, with inverse probability of treatment weighting based on propensity scores.

**Results:** Among the 41,530 patients (mean age 66.3 years; 62% male), 60.5% received DAPT. The 90-day primary outcome occurred in 10.7% of the DAPT group versus 11.6% in the monotherapy group (HR 0.82; 95% CI, 0.77–0.87). The benefit of DAPT was most pronounced when initiated within 24 hours (HR 0.74; 95% CI, 0.69–0.79). No significant benefit was observed when DAPT was initiated between 24 and 72 hours (HR 1.00; 95% CI, 0.88–1.15), and a higher risk was suggested for initiation beyond 72 hours (HR 1.25; 95% CI, 1.01–1.55). Time-dependent analysis showed benefit crossing the null at ∼42 hours.

**Conclusions:** Early initiation of DAPT—particularly within 24 hours—was associated with the greatest clinical benefit, consistent with current guideline recommendations. The therapeutic effect appeared to decline progressively beyond this period, with an estimated threshold around 42 hours.

## Introduction

Dual antiplatelet therapy (DAPT) with aspirin and clopidogrel has become a cornerstone of early secondary prevention in patients with minor ischemic stroke or high-risk transient ischemic attack (TIA).^1^ Based on the landmark trials, such as the CHANCE (Clopidogrel in High-Risk Patients With Acute Nondisabling Cerebrovascular Events), POINT trial (Platelet-Oriented Inhibition in New TIA and Minor Ischemic Stroke), and the THALES trial (The Acute Stroke or Transient Ischaemic Attack Treated with Ticagrelor and acetylsalicylic acid for Prevention of Stroke and Death)^2–4^, current guidelines recommend DAPT for patients with minor stroke or high-risk TIA who present within 24 hours of symptom onset.^5^

In real-world clinical settings, delays are common due to late symptom recognition, inter-hospital transfers, or diagnostic confirmation time.^6^ Despite these challenges, DAPT use in acute stroke management has been steadily increasing as clinicians aim to reduce early recurrence risk.^7^ However, whether starting DAPT beyond 24 hours still confers a meaningful benefit remains uncertain. Secondary analysis from the POINT and the recent INSPIRES trial (Intensive Statin and Antiplatelet Therapy for Acute High-Risk Intracranial or Extracranial Atherosclerosis) in China demonstrated that DAPT benefit remains favorable even when it is initiated up to 72 hours after symptom onset.^8,9^

Therefore, we aimed to leverage a nationwide multicenter stroke registry to assess the relationship between the time of DAPT initiation and clinical outcomes in patients with minor ischemic stroke or high-risk TIA. Our objectives were to (1) compare 90-day vascular outcomes between DAPT versus mono-antiplatelet therapy (MAPT) across different time-to-arrival intervals, and (2) estimate the time threshold for DAPT efficacy.

## Methods

### Study Design

This study analyzed data from the Clinical Research Center for Stroke-Korea-National Institute of Health (CRCS-K-NIH) registry, a multicenter, prospective, web-based registry. The study design was observational and cohort-based, incorporating consecutive patients with acute ischemic stroke or TIA. Detailed descriptions of the CRCS-K-NIH registry have been published previously.^10–12^ CRCS-K-NIH registry included patients admitted to 20 academic hospitals across South Korea between January 2011 and April 2023. The study was approved by the institutional review boards of all participating centers. A waiver of informed consent was granted owing to the use of anonymized data and minimal participant risk, with data collected for quality assessment of stroke care. Individual patient-level data are available upon reasonable request to the corresponding author, subject to legal and ethical restrictions.

### Participants

We identified patients with acute minor ischemic stroke admitted between January 2011 and April 2023, incorporating elements from the landmark minor stroke trials for high-risk TIA.^2–4^ Eligible patients were adults (≥18 years) with acute minor ischemic stroke (defined as a National Institutes of Health Stroke Scale [NIHSS] score ≤5) or high-risk TIA. High-risk TIA was defined as either the presence of an acute ischemic lesion on diffusion-weighted MRI^13^ or symptomatic intracranial arterial stenosis/occlusion.^14^ Patients had to have symptom onset within 7 days before admission and receipt of aspirin and/or clopidogrel upon admission.

We excluded patients with high risk cardioembolic source, those receiving acute reperfusion therapies (intravenous thrombolysis or endovascular thrombectomy), or antiplatelet/anticoagulant regimens other than aspirin or clopidogrel.^15^

The final study population was categorized based on the antiplatelet regimen administered upon admission: MAPT with aspirin or clopidogrel, or DAPT combining both agents.

### Outcome Measures

The primary outcome was a composite of stroke recurrence, myocardial infarction, and all-cause mortality within 90 days. Secondary outcomes included individual occurrences of recurrent stroke and all-cause mortality. Outcomes were prospectively captured during hospitalization and through structured telephone interviews and outpatient follow-ups.^10^ Outcome definitions followed standardized criteria and were consistent with prior landmark trials.^2–4^ Specifically, the definition of stroke recurrence also included cases of early neurological deterioration attributable to stroke progression, defined as worsening neurological status due to progressive ischemia, swelling of infarcted tissue, or perilesional edema on follow-up imaging.^16,17^

### Statistical Analysis

We analyzed the baseline characteristics of the entire patient cohort, regardless of the time window, summarizing continuous variables as mean and standard deviation (SD), categorical variables as frequency and proportion, and ordinal variables as median with interquartile range (IQR).

Based on current guidelines recommending DAPT initiation within 24 hours of symptom onset¹ and the INSPIRES trial allowing treatment initiation up to 72 hours,^8^ we categorized the time from symptom onset to hospital arrival into three clinically relevant intervals: within 24 hours (0–24 hours), 24 to 72 hours, and beyond 72 hours (up to 7 days).

To assess balance between treatment groups, we generated a single propensity score (PS) model, rather than separate models for each of the time interval subgroups, incorporating interaction terms between subgroup indicators and covariates.^18^ This approach was adopted to minimize the risk of subgroup inconsistency, reduced comparability, overfitting in small strata, and inadequate confounding adjustment.^18^ Using this unified PS model, we applied inverse probability of treatment weighting (IPTW) as the primary method to balance covariates, including in all subgroup analyses. Propensity score matching (PSM) was additionally performed as a secondary analysis to evaluate the robustness of findings. Covariate balance was evaluated using absolute standardized differences (ASD) before and after IPTW and PSM.

Baseline characteristics with crude, multivariable-adjusted, and propensity score–adjusted comparisons between the MAPT and DAPT groups were conducted for the overall cohort as well as within each predefined onset-to-treatment interval. For crude analyses, continuous variables were compared using the Student’s t-test, categorical variables using the Pearson chi-square test, and ordinal variables using the Wilcoxon rank sum test. For IPTW-adjusted analyses, weighted chi-square test and weighted Student’s t-test were employed, while PSM-adjusted analyses utilized conditional logistic regression models.

For primary and secondary outcomes, we plotted Kaplan-Meier curves to estimate 90-day cumulative incidence and used the log-rank test to compare DAPT vs MAPT across different time windows. Hazard ratios (HRs) with 95% confidence intervals (CIs) were estimated using Cox proportional hazards regression under three frameworks: multivariable adjustment, inverse probability of treatment weighting (IPTW), and propensity score matching (PSM). Analyses were performed for the overall cohort and stratified by prespecified onset-to-treatment intervals to assess temporal variations in DAPT efficacy. The same set of covariates—including age, sex, stroke severity (NIHSS score), body mass index, premorbid modified Rankin Scale score, Trial of Org 10172 in Acute Stroke Treatment (TOAST) classification determined by an MRI-based algorithm^15,19^, hypertension, diabetes mellitus, dyslipidemia, history of coronary artery disease, history of stroke or TIA, smoking status, symptomatic intracranial steno-occlusion, prior antiplatelet use, initial glucose level, and systolic blood pressure—was used in both the multivariable Cox model and the propensity score model. Using this unified propensity score model, we conducted IPTW-adjusted Cox regression with robust standard errors. Additionally, for validation, we performed PSM followed by Cox regression with robust standard errors to account for clustering within matched pairs.

Additionally, we conducted predefined subgroup analyses to examine whether the effect of DAPT varied according to age (<75 vs ≥75 years), sex (female vs male), arrival year (<2019 vs ≥2019, considering the implementation of clinical practice guidelines following the POINT trial), index event severity (NIHSS ≤3 vs 4–5), stenosis severity (presence vs absence of symptomatic steno-occlusion), stroke etiology (large artery atherosclerosis, small-vessel occlusion, or other causes), history of prior antiplatelet use, and history of coronary heart disease. Interaction tests were performed to assess heterogeneity across subgroups.

To examine the time-dependent effect of DAPT more granularly, we treated time from symptom onset to hospital arrival as a continuous variable. An interaction term between treatment group and time was included in the Cox model to assess time-dependent changes in HRs and identify the optimal window for DAPT. From this model, we identified the time point at which the 95% confidence interval for the hazard ratio crossed 1, indicating the loss of a statistically significant benefit of DAPT compared to MAPT. Post-hoc analyses, including subgroup analyses based on stroke etiology, symptomatic steno-occlusion, and history of antiplatelet were also conducted.

All statistical analyses were performed using SAS version 9.4 (SAS Institute Inc., Cary, NC, USA) and R version 4.4.1 (R Foundation for Statistical Computing, Vienna, Austria). A two-sided P-value of <0.05 was considered statistically significant.

### Data availability

Data are available from the corresponding author upon reasonable request.

## Results

### Baseline Characteristics and Outcomes

A total of 41,530 patients met the eligibility criteria (Figure S1), of whom 25,112 (60.5%) received DAPT and 16,418 (39.5%) received MAPT on admission. Patients selected for DAPT had more vascular risk factors than those treated with monotherapy (Table 1). The DAPT group was slightly older (67.0 vs 65.2 years) and had higher frequencies of prior stroke/TIA (20.6% vs 14.5%), coronary artery disease (8.2% vs 4.9%), and prior aspirin use (30.7% vs 18.0%). Large artery atherosclerosis (LAA) was more prevalent among DAPT patients (48.6% vs 37.2%), as was the presence of symptomatic intracranial stenosis/occlusion of the relevant artery (37.1% vs 29.4%). Baseline characteristics varied according to the time from symptom onset to hospital arrival, with differences observed in stroke mechanism and prior antiplatelet use, specifically showing an increased proportion of patients with LAA and more common prior antiplatelet use among later-arriving patients (Tables S1 to S4).

**Table 1.**
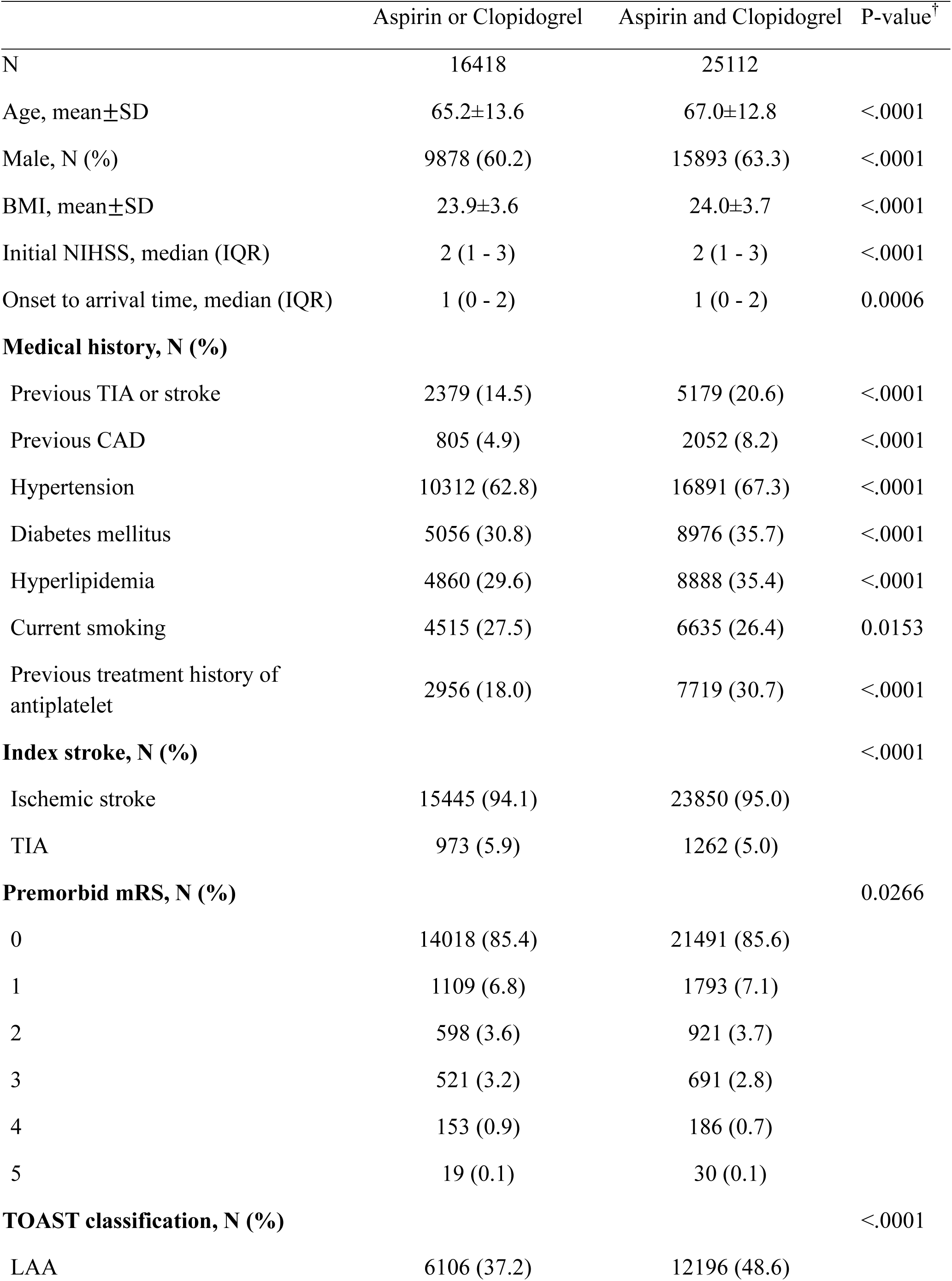
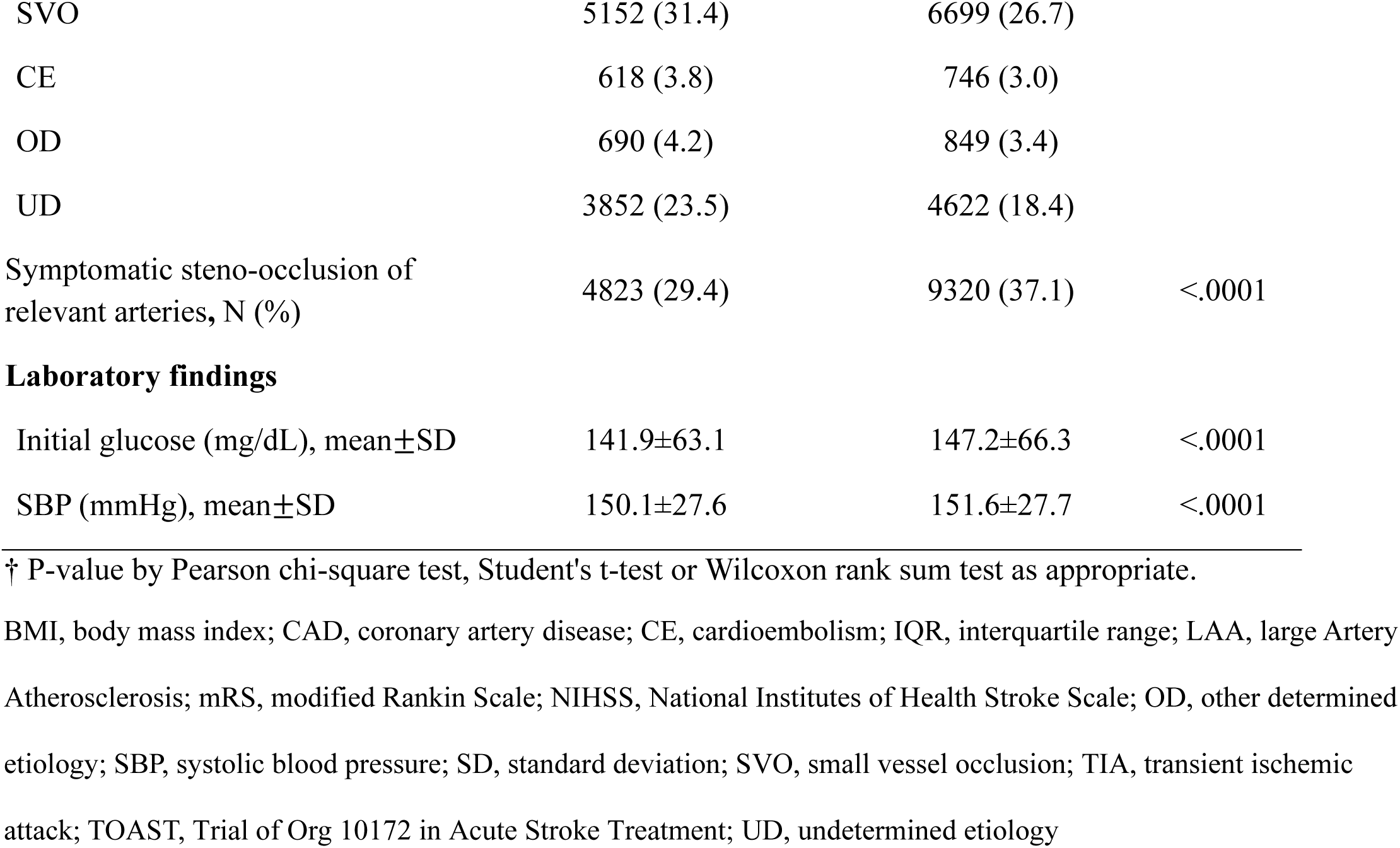
Baseline Characteristics of Patients in the Whole Time Window.

|  | Aspirin or Clopidogrel | Aspirin and Clopidogrel | P-value<sup>†</sup> |
| --- | --- | --- | --- |
| N | 16418 | 25112 |  |
| Age, mean±SD | 65.2±13.6 | 67.0±12.8 | <.0001 |
| Male, N (%) | 9878 (60.2) | 15893 (63.3) | <.0001 |
| BMI, mean±SD | 23.9±3.6 | 24.0±3.7 | <.0001 |
| Initial NIHSS, median (IQR) | 2 (1 - 3) | 2 (1 - 3) | <.0001 |
| Onset to arrival time, median (IQR) | 1 (0 - 2) | 1 (0 - 2) | 0.0006 |
| **Medical history, N (%)** |  |  |  |
| Previous TIA or stroke | 2379 (14.5) | 5179 (20.6) | <.0001 |
| Previous CAD | 805 (4.9) | 2052 (8.2) | <.0001 |
| Hypertension | 10312 (62.8) | 16891 (67.3) | <.0001 |
| Diabetes mellitus | 5056 (30.8) | 8976 (35.7) | <.0001 |
| Hyperlipidemia | 4860 (29.6) | 8888 (35.4) | <.0001 |
| Current smoking | 4515 (27.5) | 6635 (26.4) | 0.0153 |
| Previous treatment history of antiplatelet | 2956 (18.0) | 7719 (30.7) | <.0001 |
| **Index stroke, N (%)** |  |  | <.0001 |
| Ischemic stroke | 15445 (94.1) | 23850 (95.0) |  |
| TIA | 973 (5.9) | 1262 (5.0) |  |
| **Premorbid mRS, N (%)** |  |  | 0.0266 |
| 0 | 14018 (85.4) | 21491 (85.6) |  |
| 1 | 1109 (6.8) | 1793 (7.1) |  |
| 2 | 598 (3.6) | 921 (3.7) |  |
| 3 | 521 (3.2) | 691 (2.8) |  |
| 4 | 153 (0.9) | 186 (0.7) |  |
| 5 | 19 (0.1) | 30 (0.1) |  |
| **TOAST classification, N (%)** |  |  | <.0001 |
| LAA | 6106 (37.2) | 12196 (48.6) |  |
| SVO | 5152 (31.4) | 6699 (26.7) |  |
| CE | 618 (3.8) | 746 (3.0) |  |
| OD | 690 (4.2) | 849 (3.4) |  |
| UD | 3852 (23.5) | 4622 (18.4) |  |
| Symptomatic steno-occlusion of relevant arteries, N (%) | 4823 (29.4) | 9320 (37.1) | <.0001 |
| Laboratory findings |  |  |  |
| Initial glucose (mg/dL), mean±SD | 141.9±63.1 | 147.2±66.3 | <.0001 |
| SBP (mmHg), mean±SD | 150.1±27.6 | 151.6±27.7 | <.0001 |

† P-value by Pearson chi-square test, Student's t-test or Wilcoxon rank sum test as appropriate.
BMI, body mass index; CAD, coronary artery disease; CE, cardioembolism; IQR, interquartile range; LAA, large Artery Atherosclerosis; mRS, modified Rankin Scale; NIHSS, National Institutes of Health Stroke Scale; OD, other determined etiology; SBP, systolic blood pressure; SD, standard deviation; SVO, small vessel occlusion; TIA, transient ischemic attack; TOAST, Trial of Org 10172 in Acute Stroke Treatment; UD, undetermined etiology

The median follow-up duration was 367 days (IQR, 353–381) while a total of 94.4% of patients were followed up for at least 90 days. During the follow-up period, 6,249 patients (15.0%) experienced recurrent stroke, 74 (0.2%) had an acute myocardial infarction, and 2,325 (5.6%) died from any cause. Event rates of the primary outcome were slightly lower in the DAPT group (10.7% vs 11.6%, Figure 1A). Recurrent stroke occurred in 10.0% of patients who received DAPT, compared to 11.0% with MAPT (Figure S2A). All-cause mortality by 90 days was low and similar between groups (1.40% DAPT vs 1.44% MAPT, Figure S3A).

**Figure 1.**
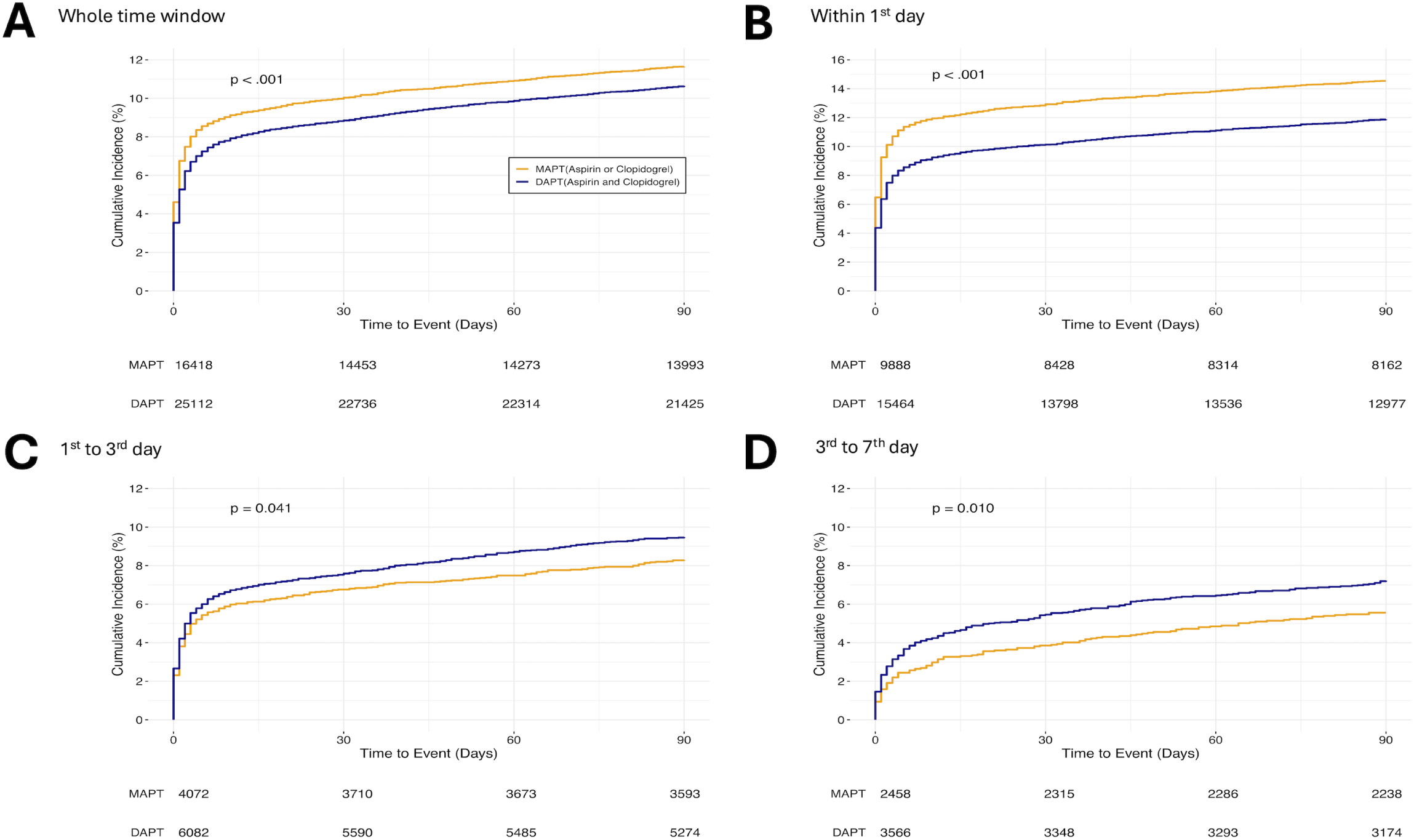
Cumulative incidence of the primary outcome (composite of stroke recurrence, myocardial infarction, and all-cause mortality) up to 3 months after symptom onset by antiplatelet use (aspirin or clopidogrel vs. aspirin and clopidogrel) and stratified by four time windows. (A) Whole time window (0–90 days) (B) Within 1st day (0–1 day) (C) 1st to 3rd day (1–3 days) (D) 3rd to 7th day (3–7 days) The number at risk for each group is displayed below the x-axis. P-values were calculated using the log-rank test.

### Time-Dependent effect of DAPT over MAPT

After propensity score weighting and matching, baseline covariates were well balanced between groups (ASD <0.1 for key variables), confirming the effectiveness of adjustment strategies (Tables S1–S4 and Figure S4). DAPT was consistently associated with lower risks of the primary composite outcome and stroke recurrence across all analytic models (multivariable adjustment, IPTW, PSM). HRs for the primary outcome ranged from 0.82 to 0.85, and for stroke recurrence from 0.80 to 0.83. Mortality rates at 90 days were similar between groups (HRs 0.90– 0.92; Table 2).

**Table 2.** Adjusted Hazard Ratios for Primary and Secondary Outcomes by Time Window (x : Duration from onset to arrival (in days))

| Time window | Whole time window |  | 0≤x≤1 |  | 1<x≤3 |  | 3<x≤7 |  | P for interaction |
| --- | --- | --- | --- | --- | --- | --- | --- | --- | --- |
|  | Adjusted HR (95% CI) | P value | Adjusted HR (95% CI) | P value | Adjusted HR (95% CI) | P value | Adjusted HR (95% CI) | P value |  |
| Primary outcome | 0.82 (0.77-0.87) | <.0001 | 0.74 (0.69-0.79) | <.0001 | 1.04 (0.91-1.19) | 0.5632 | 1.19 (0.96-1.46) | 0.1102 | <.0001 |
| Secondary outcomes |  |  |  |  |  |  |  |  |  |
| Stroke recurrence | 0.80 (0.75-0.85) | <.0001 | 0.73 (0.68-0.78) | <.0001 | 1.03 (0.90-1.19) | 0.6650 | 1.13 (0.90-1.40) | 0.2945 | <.0001 |
| All-cause death | 0.92 (0.77-1.09) | 0.3209 | 0.77 (0.62-0.96) | 0.0190 | 1.04 (0.74-1.46) | 0.8360 | 1.55 (0.97-2.45) | 0.0643 | 0.0190 |
| After IPTW | HR (95% CI) | P value | HR (95% CI) | P value | HR (95% CI) | P value | HR (95% CI) | P value |  |
| Primary outcome | 0.82 (0.77-0.87) | <.0001 | 0.74 (0.69-0.79) | <.0001 | 1.00 (0.88-1.15) | 0.9448 | 1.25 (1.01-1.55) | 0.0409 | <.0001 |
| Secondary outcomes |  |  |  |  |  |  |  |  |  |
| Stroke recurrence | 0.80 (0.75-0.85) | <.0001 | 0.72 (0.67-0.78) | <.0001 | 1.00 (0.87-1.16) | 0.9927 | 1.17 (0.93-1.46) | 0.1793 | <.0001 |
| All-cause death | 0.90 (0.75-1.06) | 0.2107 | 0.77 (0.62-0.95) | 0.0173 | 0.95 (0.67-1.35) | 0.7827 | 1.61 (1.00-2.61) | 0.051 | 0.0201 |
| After PSM | HR (95% CI) | P value | HR (95% CI) | P value | HR (95% CI) | P value | HR (95% CI) | P value |  |
| Primary outcome | 0.85 (0.79-0.90) | <.0001 | 0.77 (0.71-0.83) | <.0001 | 1.04 (0.89-1.21) | 0.6337 | 1.21 (0.95-1.52) | 0.1184 | <.0001 |
| Secondary outcomes |  |  |  |  |  |  |  |  |  |
| Stroke recurrence | 0.83 (0.77-0.89) | <.0001 | 0.76 (0.70-0.82) | <.0001 | 1.03 (0.88-1.21) | 0.7316 | 1.15 (0.90-1.47) | 0.2705 | <.0001 |
| All-cause death | 0.91 (0.76-1.10) | 0.3401 | 0.80 (0.63-1.01) | 0.0653 | 0.99 (0.67-1.46) | 0.9564 | 1.41 (0.85-2.34) | 0.1785 | 0.1183 |

Hazard ratios were derived from Cox proportional hazards regression models. The adjusted model included the same covariates as those used in the propensity score model. In the IPTW analysis, a weighted Cox model with robust standard errors was employed. In the PSM analysis, robust standard errors were used to account for clustering within matched pairs. CI, confidence interval; HR, hazard ratio; IPTW, inverse probability of treatment weighting ; PSM, propensity score matching

#### Within 24 hours of onset

DAPT initiated within 24 hours was associated with significantly lower 90-day event rates compared to monotherapy (11.9% vs 14.5%, Figure 1B), primarily due to reduced stroke recurrence (11.3% vs 14.0%) and a slight reduction in mortality (1.3% vs 1.6%). The benefit was consistent across all models: HR 0.74 (95% CI, 0.69–0.79) by multivariable and IPTW adjustment; HR 0.77 (95% CI, 0.71–0.83) by PSM (Table 2).

#### Between 24 and 72 hours

Event rates were comparable between groups (primary outcome: 9.5% vs 8.3%, Figure 1C; recurrent stroke: 8.6% vs 7.5%, Figure S2C; mortality: 1.5% vs 1.4%, Figure S3C). Adjusted HRs showed no significant difference: HR 1.00–1.04 across all models (Table 2).

#### Between 72 hours and 7 days

DAPT was associated with higher event rates than MAPT (primary outcome: 7.2% vs 5.6%, Figure 1D; stroke recurrence: 6.3% vs 5.1%, Figure S2D; mortality: 1.7% vs 1.1%, Figure S3D). HRs indicated a potential excess risk with DAPT: HR 1.25 (95% CI, 1.01–1.55) in IPTW; similar trends in multivariable (HR 1.19) and PSM (HR 1.21) models (Table 2).

To explore whether the treatment effect of DAPT varied across subgroups, we conducted predefined subgroup analyses (Figure 2). While DAPT was generally associated with a reduced risk of recurrent vascular events, there was notable heterogeneity across clinical subgroups, particularly among patients who arrived >3 days after symptom onset, in whom the treatment effect reversed — favoring MAPT in those with LAA or prior antiplatelet use.

**Figure 2.**
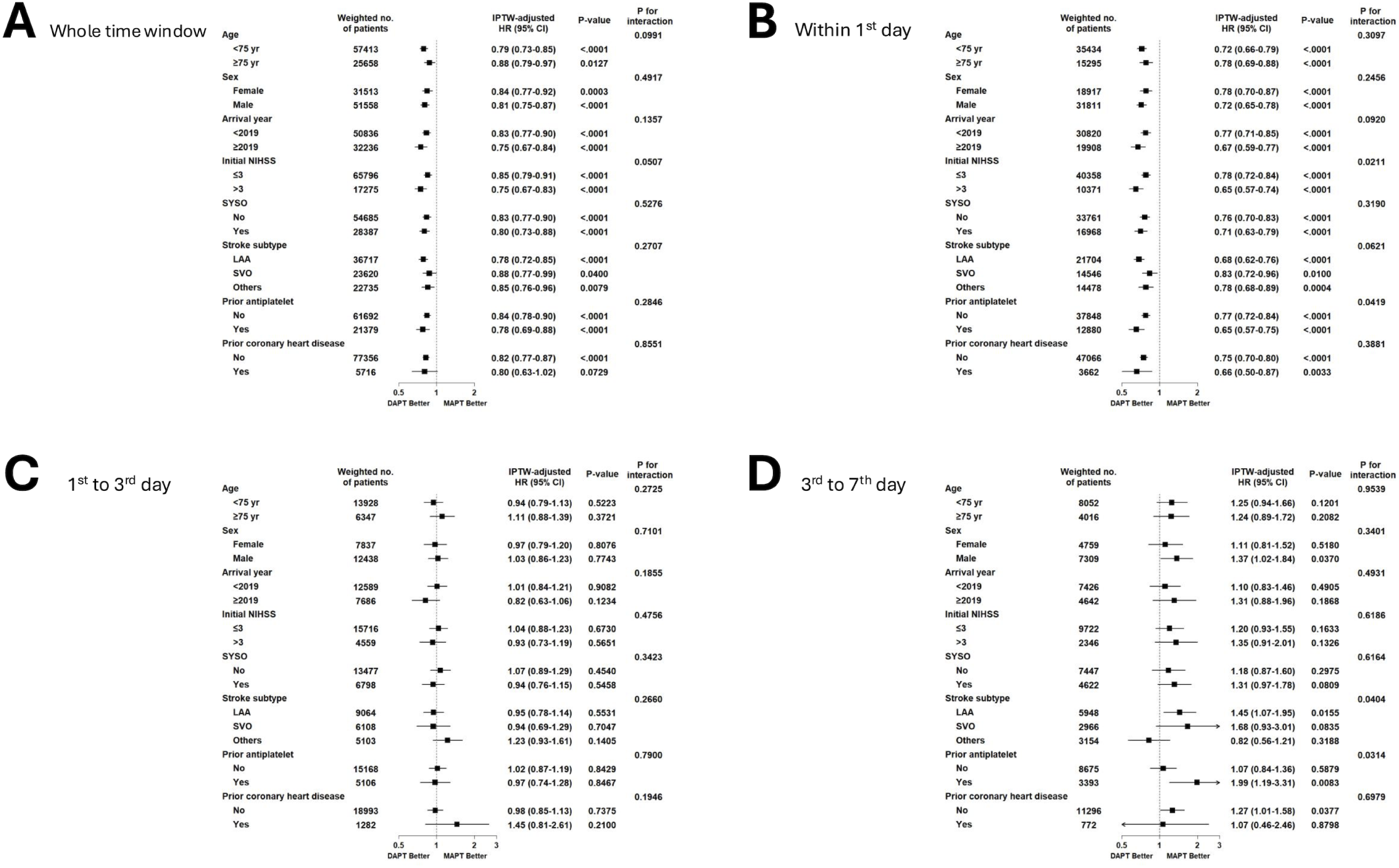
Hazard ratios for the primary efficacy outcome across prespecified subgroups. (A) Whole time window (0–90 days) (B) Within 1st day (0–1 day) (C) 1st to 3rd day (1–3 days) (D) 3rd to 7th day (3–7 days) The weighted number of patients in each subgroup is presented. P-values indicate interaction effects between treatment and subgroup characteristics. DAPT indicates dual antiplatelet therapy; HR, hazard ratio; IPTW, inverse probability of treatment weighting; LAA, large artery atherosclerosis; MAPT, mono antiplatelet therapy; NIHSS, National Institutes of Health Stroke Scale; SVO, small vessel occlusion; SYSO, symptomatic steno-occlusion of the relevant artery.

In analyses treating time to treatment as a continuous variable, we estimated the threshold beyond which DAPT no longer conferred a statistically significant benefit compared to MAPT. The estimated thresholds were approximately 42 hours for the primary composite outcome, 45 hours for stroke recurrence, and 17 hours for all-cause mortality (Figure 3). Post-hoc subgroup analyses revealed variations in the estimated optimal timing: 16 hours for small vessel occlusion, 39 hours for cardioembolism/other determined etiology/undermined etiology, and 41 hours for LAA (Figure S5). The thresholds were also shorter among patients without symptomatic steno-occlusion (37 vs 44 hours; Figure S6) and those with prior antiplatelet use (37 vs 41 hours; Figure S7).

**Figure 3.**
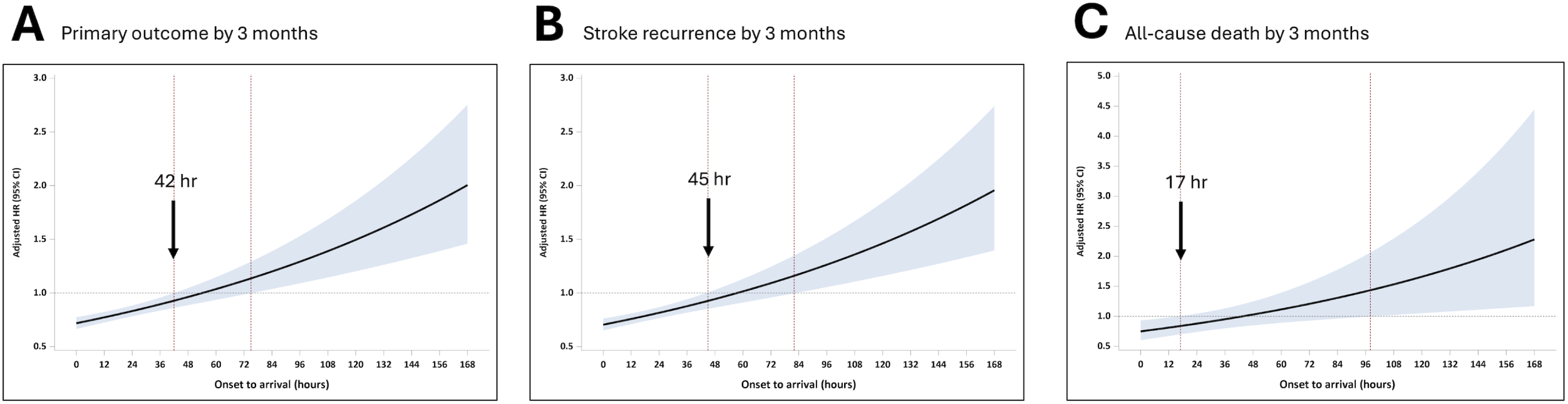
Adjusted hazard ratios for the primary outcome over time. Adjusted hazard ratio (HR) curves estimated using a multivariable Cox proportional hazards model, with time from symptom onset to hospital arrival modeled as a continuous variable. The black line represents the HR, and the shaded area indicates the 95% confidence interval. Red arrows indicate the upper bound of the 95% confidence interval at the estimated time points where the HR crosses 1.0, suggesting potential changes in risk. (A) Primary outcome at 3 months (B) Stroke recurrence at 3 months (C) All-cause mortality at 3 months

## Discussion

In this nationally representative, real-world cohort of patients with minor stroke or high-risk TIA, early DAPT initiation—especially within the first 24 hours—was consistently associated with improved outcomes in reducing recurrent ischemic events, reinforcing the urgency emphasized in clinical guidelines.^5^ However, beyond this period, the benefit became less clear. Time-continuous Cox analyses suggested that the therapeutic window for DAPT may extend up to approximately 42 to 45 hours, supporting the notion that DAPT may be most effective during the early phase when unstable thrombi or active plaques are biologically active.^20^

These findings extend prior trial evidence and offer insight into the optimal therapeutic window for DAPT. All three trials—CHANCE, POINT, and THALES trials—emphasized treatment initiation within 12–24 hours of symptom onset, aligning with our observed benefit in the 0–24-hour window.^2–4^ In patients treated within 24 hours, DAPT conferred a 2.6% absolute stroke risk reduction, closely aligning with CHANCE (3.5%) and POINT (1.5%).^3,4^ THALES further validated the early-treatment principle (though using ticagrelor instead of clopidogrel) by showing a 17% relative risk reduction in stroke at 30 days.^2^ In line with these trials, our study’s results reinforce the critical importance of early initiation – the sooner, the better.

A key novel contribution of our analysis is the estimation of a real-world threshold— approximately 42 hours after symptom onset—beyond which the benefit of DAPT diminished. Although the greatest benefit was seen with initiation within the first 24 hours, our findings suggest that the therapeutic window may extend beyond this period. A post hoc analysis of the POINT trial demonstrated that the benefit of DAPT remained favorable when initiated up to 72 hours after symptom onset, with the 95% confidence interval for major ischemic events crossing above 1 at around 48 hours — aligning with our findings of diminishing benefit beyond this timeframe.^9^ Antiplatelet Therapy in Acute Mild to Moderate Ischemic Stroke trial (ATAMIS) also showed DAPT reduced END within 48hr of symptom onset.^21^

A recent trial, INSPIRES, which focused on symptomatic intracranial or extracranial atherosclerosis and those with multiple acute infarctions, found that initiating DAPT up to 72 hours after stroke onset reduced 90-day stroke risk compared with aspirin alone (1.9% absolute risk reduction), without excess severe bleeding.^8^ Our findings are generally consistent with this result but suggest a somewhat narrower therapeutic window. Specifically, in patients with LAA stroke, the estimated threshold for DAPT benefit appeared to be around 42 hours after symptom onset. Several factors may explain this shorter optimal window compared to INSPIRES. Patients enrolled in clinical trials are typically selected based on strict inclusion criteria, tend to be healthier, more homogeneous, and more adherent to treatment protocols.^22^ In contrast, real-world populations are more heterogeneous, with greater variability in comorbidities, treatment adherence, and stroke mechanisms, which may have led to an earlier attenuation of DAPT benefit in our study.

In our study, time-to-treatment was defined as the interval from symptom onset to hospital arrival, serving as a pragmatic proxy for DAPT initiation in actual clinical workflows, where delays from triage, imaging, and consent are common.^23^ In contrast, prior randomized trials used symptom onset to randomization as the inclusion window, which inherently includes delays for diagnostic evaluations, consent, and randomization procedures.^2–4,8^ For example, the INSPIRES trial allowed enrollment up to 72 hours post-onset, with medications administered within 1 hour after randomization.^8^ Notably, the median onset-to-randomization time in INSPIRES was around 46 hours, suggesting that treatment often began later in the course of stroke evolution.^24^ While direct comparisons are limited due to differing definitions and study designs, the estimated 42-hour threshold observed in our study may reflect a broadly similar therapeutic window, highlighting the need for further research to refine optimal timing for DAPT initiation in real-world clinical practice.

Several limitations of this study should be acknowledged. First, as a secondary analysis of a multicenter, prospective cohort study, baseline differences existed between treatment groups. Although we adjusted for known confounders using propensity methods, unmeasured factors— such as clinicians’ judgment of plaque instability or patient frailty—could have influenced both treatment timing and outcomes. Second, major bleeding events were not systematically captured in our registry. However, prior major trials of early DAPT (CHANCE, POINT, THALES, INSPIRES) consistently reported low rates of severe bleeding (approximately 0.3% to 0.9%), supporting the overall safety of short-term DAPT in this setting.^2–4,8^ Third, data on antiplatelet dosing and loading protocols were not standardized in our registry. This limitation, also noted in prior real-world studies, reflects the variability in physician-driven treatment decisions.^25^ Fourth, most participating centers were academic hospitals, which may limit generalizability.

Nevertheless, the alignment of our results with national stroke audit data suggests broader applicability.^26^ Finally, the study was conducted in South Korea, with a predominantly East Asian population and healthcare system, which may limit generalizability to non–East Asian populations or health systems. However, the consistency of these results with randomized trials strengthens their external validity.^2–4,8^

## Conclusion

This study provides real-world evidence that DAPT initiated within 24 hours after a minor ischemic stroke or high-risk TIA confers significant clinical benefit, consistent with findings from CHANCE, POINT, and THALES trials. Importantly, our data suggest that therapeutic benefit may extend up to approximately 42 hours, potentially offering an extended window for intervention in patients unable to receive immediate treatment. Prospective studies are warranted to validate these observations and to inform guidelines on optimal DAPT initiation timing for secondary stroke prevention.

## Nonstandard Abbreviations and Acronyms

ATAMIS: Antiplatelet Therapy in Acute Mild to Moderate Ischemic Stroke
CE: cardioembolism
CHANCE: Clopidogrel in High-Risk Patients With Acute Nondisabling Cerebrovascular Events
CRCS-K-NIH: Clinical Research Center for Stroke-Korea-National Institute of Health
DAPT: dual antiplatelet therapy
DWI: diffusion-weighted imaging
END: early neurological deterioration
INSPIRES.: Intensive Statin and Antiplatelet Therapy for Acute High-Risk Intracranial or Extracranial Atherosclerosis
LAA.: large artery atherosclerosis
MAPT: mono-antiplatelet therapy
mRS: modified Rankin Scale
NIHSS: National Institutes of Health Stroke Scale
POINT: Platelet-Oriented Inhibition in New TIA and Minor Ischemic Stroke
SVO: small vessel occlusion
THALES: The Acute Stroke or Transient Ischaemic Attack Treated with Ticagrelor and Acetylsalicylic Acid for Prevention of Stroke and Death
TOAST: Trial of Org 10172 in Acute Stroke Treatment

